# Reduced access to care among older American adults during CoVID-19 pandemic: results from a prospective cohort study

**DOI:** 10.1101/2020.11.29.20240317

**Authors:** Ashis Kumar Das, Devi Kalyan Mishra, Saji Saraswathy Gopalan

**Affiliations:** Research Group, The World Bank, Washington DC, USA; Department of Community Medicine, Hitech Medical College, Rourkela, India; Human Development Department, The World Bank, Washington DC, USA

## Abstract

**Background:** Due to preexisting conditions, older adults are at higher risk of COVID-19 related severe complications. Current evidence is limited on access to care for older adults during the COVID-19 pandemic.

**Objectives:** To examine the extent of reduced access to care among older American adults during the COVID-19 pandemic, identify predictors and reasons of reduced access.

**Materials and methods:** Using publicly available data from the COVID-19 module (interim release) of the Health and Retirement Study, we undertook descriptive analyses of older adults stratified by sex, age group, race, education, marital status, employment, receipt of social security benefits, health insurance, number of limitations in activities of daily living and pre-existing conditions. Associations between reduced access to care and predictors were estimated using a multivariable logistic regression model.

**Results:** About 30% of respondents delayed or avoided care during the pandemic. Reduced access was more likely to be reported by respondents that were female, younger, educated, not receiving social security benefits, with limitations in daily activities and three preexisting conditions. In terms of the reasons, the majority of the respondents (45.9%) reported that their visit was either cancelled or rescheduled by the provider; 13.9% thought they could wait, 10.9% could not get an appointment, 9.1% found it unaffordable, and 7.4% were afraid to visit the provider. Respondents reported of reduced access to doctor’s visits, surgery, prescription filling, and dental care.

**Conclusions:** We suggest urgent attention on improving access to care for older adults during the pandemic. For nonemergency conditions and routine care that can be delivered virtually, telehealth services can be strengthened. Additionally, health messaging can reemphasize that neglecting medical care might lead to increased morbidity and mortality among older adults from preexisting illnesses.

## 1. Introduction

As of November 24, 2020, the US has reported approximately 12.3 million confirmed cases and 257,016 deaths due to COVID-19.^1^ The global evidence says that a pandemic of this scale has certain potential indirect effects on essential and routine healthcare usage patterns.^2–4^ There could be several plausible determinants driving the access to essential healthcare patterns, apart from the pandemic mitigation strategies such as lock down and social distancing, and severity of the pandemic itself. The existing social, demographic, and economic contexts, and underlying health conditions could be a few of such determinants. However, the current evidence is limited globally and specifically, in the USA on the indirect effects of COVID-19 on access to care for older adults during the pandemic. Due to preexisting conditions, older adults are usually at higher risk of COVID-19 related severe complications.^5^ Additionally, negligence of routine medical care for chronic illnesses can lead to higher morbidity and mortality among older adults. There are several attempts to inform the potential impact of COVID-19 on emergency care, cardiovascular disease care, cancer care, and other essential outpatient care encompassing the general population.^4,6–10^ However, to the best of our knowledge no study has explored the dimensions of access to care among the elderly population. Understanding the neglect of healthcare and potential reasons among older adults could be relevant for the policy makers to improve elderly health as the pandemic continues.

In this context, we explored access to care and its determinants among older American adults during the pandemic using a nationally representative survey. We also assessed the reasons for reduced access to care.

## 2. Materials and methods

### 2.1 Data source

The data were obtained from the COVID-19 module (interim release) of the Health and Retirement Study (HRS). The HRS is an ongoing prospective cohort study that is nationally representative of adults ages 50 and over in the USA.^11^ Participants from the HRS are interviewed every two years and are followed-up actively through regular mail contact and phone interviews. The COVID-19 module of HRS 2020 was administered to half of the random subsample of households who were originally assigned to enhanced face-to-face interviewing. This was done over two phases using two randomly halved subsamples during June and September. In this study, we use the interim release data from the June study that includes 3,266 respondents. After excluding those with missing values, 3,129 respondents were included in the final analysis.

### 2.2 Variables

The outcome of interest was reduced access to health care during the pandemic. In the COVID-19 module, the respondents were asked if they delayed getting medical care or did not get at all since March 2020. This response was collected as a binary variable in the data – yes if they delayed or forgone medical care and otherwise no. The predictors were age in groups (50-59, 60-69, 70-79, 80 and above), sex, education (below high school, high school, college and post-college), employment (employed, unemployed, retired, homemaker), marital status (legally married and living with spouse, others), health insurance (none, Medicare/Medicaid, private, TriCare/VA), receipt of social security benefits, number of difficulties in activities of daily living – ADL (none, at least 1), and number of self-reported pre-existing conditions (none, 1, 2, 3, 4 or more). Difficulties in caring for the self as a result of health or physical issues constitute ADLs. Caring for the self includes activities such as bathing, dressing up, eating, getting in or out of bed, or using toilets. Pre-existing conditions consisted of hypertension, diabetes, cancer, chronic lung disease, heart conditions, stroke, psychiatric problems, and arthritis.

### 2.3 Statistical methods

We undertook descriptive analyses for the respondent characteristics and presented the results stratified by subgroups for each characteristic. Correlation was tested among all respondent characteristics with the Pearson’s correlation coefficient. Associations between reduced access to care and predictors (age group, sex, education, employment, marital status, health insurance, receipt of social security benefits, number of ADLs, and number of preexisting conditions) were estimated using a multivariable logistic regression model adjusting for all predictors. We considered the associations statistically significant if the p-value was below 0.05. The statistical analyses were performed using Stata Version 15 (StataCorp LLC. College Station, TX). Sampling weights were applied to all analyses to account for the HRS sampling design.

## 3. Results

### 3.1 Descriptive analysis

The profile of respondents is shown in table 1. Out of 3,129 respondents, slightly more than half (53.1%) were females with the single largest age group being 60 to 69 years (38.9%). Whites constituted 79.5 per cent, followed by Blacks (10.9%) and others (9.6%). Around 41 per cent were legally married and living with their spouses, 44 per cent had college education, and about three-fifths received Social Security benefits (60.7%). A little above half (54.3%) had Medicare/Medicaid insurance, and a vast majority (88.4%) did not have any limitations in activities of daily living. Very few (14.4%) did not have any preexisting conditions. Closer to 30 per cent respondents reported of reduced access to care.

**Table 1.** Sample characteristics.

| **Variable** | **n** | **%** |
| --- | --- | --- |
| **Sex** |  |  |
| Female | 1,859 | 53.1 |
| Male | 1,270 | 46.9 |
| **Age group (Years)** |  |  |
| 50–59 | 650 | 20.9 |
| 60–69 | 1,109 | 38.9 |
| 70–79 | 728 | 26.0 |
| 80 and older | 642 | 14.3 |
| **Race** |  |  |
| Black | 664 | 10.9 |
| White | 2,103 | 79.5 |
| Others | 362 | 9.6 |
| **Education** |  |  |
| Below HS | 227 | 5.4 |
| High school | 1,203 | 36.0 |
| College | 1,283 | 44.0 |
| Post college | 416 | 14.6 |
| **Marital status** |  |  |
| Legally married and living with spouse | 1,434 | 41.4 |
| Others | 1,695 | 58.6 |
| **Employment** |  |  |
| Employed | 862 | 31.3 |
| Unemployed | 630 | 18.4 |
| Retired | 1,475 | 46.0 |
| Homemaker | 162 | 4.4 |
| **Receipt of social security benefits** |  |  |
| Yes | 1,122 | 39.3 |
| No | 2,007 | 60.7 |
| **Health insurance** |  |  |
| No insurance | 160 | 4.5 |
| Medicare/Medicaid | 1,816 | 54.3 |
| Private/Employer | 984 | 35.7 |
| TriCare/VA | 169 | 5.5 |
| **Number of ADLs** |  |  |
| 0 | 2,720 | 88.4 |
| At least 1 | 409 | 11.6 |
| **Number of pre-existing conditions** |  |  |
| 0 | 397 | 14.4 |
| 1 | 697 | 22.7 |
| 2 | 801 | 26.7 |
| 3 | 652 | 19.3 |
| 4 or more | 582 | 17.0 |
| **Reduced access to care** |  |  |
| Yes | 936 | 30.2 |
| No | 2,193 | 69.8 |
| Total | 3,129 | 100 |

**Table 2.** Predictors of reduced access to care (N=936)

| **Variable** |  | **Reduced access (%)** | **Adjusted odds ratio** | **95% confidence interval** | **p value** |
| --- | --- | --- | --- | --- | --- |
| **Sex** |  |  |  |  |  |
|  | Female | 32.4 | Reference |  |  |
|  | Male | 27.8 | 0.78 | 0.61-0.98 | 0.033 |
| **Age group (Years)** |  |  |  |  |  |
|  | 50-59 | 37.1 | Reference |  |  |
|  | 60-69 | 33.3 | 0.86 | 0.62-1.20 | 0.388 |
|  | 70-79 | 26.0 | 0.69 | 0.45-1.05 | 0.084 |
|  | 80 and older | 19.5 | 0.43 | 0.27-0.66 | <0.001 |
| **Race** |  |  |  |  |  |
|  | Black | 30.5 | Reference |  |  |
|  | White | 30.6 | 1.09 | 0.83-1.44 | 0.531 |
|  | Others | 27.0 | 0.80 | 0.52-1.24 | 0.32 |
| **Education** |  |  |  |  |  |
|  | Below HS | 17.1 | Reference |  |  |
|  | High school | 26.6 | 1.67 | 1.01-2.76 | 0.047 |
|  | College | 31.7 | 2.04 | 1.23-3.37 | 0.005 |
|  | Post college | 39.7 | 3.05 | 1.75-5.30 | <0.001 |
| **Marital status** |  |  |  |  |  |
|  | Legally married and living with spouse | 31.3 | Reference |  |  |
|  | Others | 29.4 | 0.86 | 0.68-1.08 | 0.198 |
| **Employment** |  |  |  |  |  |
|  | Employed | 35.7 | Reference |  |  |
|  | Unemployed | 31.3 | 0.85 | 0.59-1.23 | 0.392 |
|  | Retired | 27.1 | 1.02 | 0.73-1.42 | 0.906 |
|  | Homemaker | 19.6 | 0.56 | 0.31-1.02 | 0.057 |
| **Receipt of social security benefits** |  |  |  |  |  |
|  | No | 36.9 | Reference |  |  |
|  | Yes | 25.9 | 0.65 | 0.46-0.93 | 0.018 |
| **Health insurance** |  |  |  |  |  |
|  | No insurance | 30.8 | Reference |  |  |
|  | Medicare/Medicaid | 27.5 | 0.99 | 0.57-1.73 | 0.983 |
|  | Private | 34.7 | 0.96 | 0.56-1.64 | 0.884 |
|  | TriCare/VA | 27.4 | 0.97 | 0.47-2.00 | 0.938 |
| **Number of ADLs** |  |  |  |  |  |
|  | 0 | 28.9 |  |  |  |
|  | At least 1 | 40.6 | 2.25 | 1.64-3.09 | <0.001 |
| **Number of pre-existing conditions** |  |  |  |  |  |
|  | 0 | 27.4 | Reference |  |  |
|  | 1 | 31.0 | 1.39 | 0.92-2.11 | 0.12 |
|  | 2 | 31.7 | 1.44 | 0.96-2.17 | 0.082 |
|  | 3 | 32.5 | 1.72 | 1.11-2.68 | 0.016 |
|  | 4 or more | 26.6 | 1.26 | 0.80-1.98 | 0.31 |
|  | Total | 30.2 |  |  |  |

### 3.2 Predictors of reduced access to care

There were not strong correlations between predictors and the Pearson’s correlation coefficients ranged from -0.27 to 0.6. Significantly less males than females (adjusted odds ratio 0.78; p value 0.033) and older respondents than the younger ones (aOR 0.43; p value <0.001) reported reduced access to care. Similarly, respondents receiving social security benefits (aOR 0.65; p value 0.018) were less likely to have reduced access to care. Compared to respondents educated below high school, educated respondents were more likely to experience reduced access to care – high school (aOR 1.67; p value 0.047), college (aOR 2.04; p value 0.005) and post-college (aOR 3.05; p value <0.001). Likewise, respondents with limitations in activities of daily living (aOR 2.25; p value <0.001) and those with three preexisting conditions (aOR 1.72; p value 0.016) were more likely to report reduced access. Race, marital status, employment, and health insurance were not associated with reduced access.

When asked about the reasons for reduced access, the majority of the respondents reported their visit was either cancelled or rescheduled by the provider (45.9%), whereas 13.9 per cent thought they could wait, 10.9 per cent could not get an appointment, 9.1 per cent found it unaffordable, and 7.4 per cent were afraid to visit the provider. There were 12.8 per cent with other reasons for reduced access. Among the various components of health care that were either delayed or avoided, 58.8 per cent of respondents did so for visiting a doctor, 14 per cent for surgery, 4.9 per cent for filling prescriptions, and 76.5 per cent for dental care. It is worth noting that respondents reported reduced access to multiple components of care simultaneously. Therefore, the proportions may add up to more than 100 per cent.

## 4. Discussion

The COVID-19 pandemic has directly and indirectly affected access to health care across the world.^7,8,12–14^ In this study, we show that 30 per cent of older adults either delayed or forgone care during the pandemic. Moreover, reduced access was more likely to be reported by respondents that were female, younger, educated, not receiving social security benefits, with limitations in daily activities and three preexisting conditions.

In a web-based survey administered to American adults, the prevalence of reduced access among older adults over 65 years was 33.5 per cent.^15^ This study also found females, multiple preexisting medical conditions, higher education and having health insurance were significantly associated with reduced access among all age groups. The Research and Development Survey (RANDS) undertaken by the National Center for Health Statistics during COVID-19 shows 39.5 per cent of older adults had reduced access to care.^16^ Overall, females and education of above bachelor’s degree were more likely to report missing care. Additionally, about 31 per cent of older adults reported to have scheduled at least one telehealth appointment. The proportion of respondents reporting reduced access to care in these studies are higher than our study. The differences in the reduced access to care are possibly due to differing methodologies and recall periods. For instance, our study includes a nationally representative sample of older adults, whereas it was not the case with other studies. However, in line with these similar studies, our study finds female respondents and those with higher education levels tend to report reduced access to care during the pandemic. Among the elderly, research shows that being female is associated with reduced access even during non-pandemic times.^17,18^ We believe that respondents with higher education could be more aware of the risk of exposure to COVID-19 infection and hence delayed or avoided care altogether.^19^

As reported by the respondents in our study, the primary reasons for reduced access were due to cancellation, rescheduling, or not getting appointments. During the pandemic, health care providers and practices are striving to maintain stricter infection prevention and control guidelines with limited resources.^20,21^ This could have led to cancellations and rescheduling of visits to keep the patients and providers safe from potential spread of the infection. Respondents that need regular medical visits or prescription filling are more likely to have reduced access during the pandemic. Ironically, most of them suffer from preexisting conditions and research shows that they are at risk of developing serious complications if infected with COVID-19.^5^ Based on our findings, we suggest urgent attention on improving access to care for older adults during the pandemic. For nonemergency conditions and routine care that can be delivered virtually, strengthening telehealth services will be helpful. Additionally, older adults’ awareness on the availability and benefits of telehealth can be improved. Older adults would have to be reassured that the providers and healthcare facilities are taking adequate measures to ensure a safe environment and neglecting medical care might lead to increase morbidity and mortality from pre-existing illnesses.^22^ Improving involvement of medical volunteers and voluntary organizations in supporting older adults can ensure timely care.

Our study has three limitations. First, the HRS information is self-reported and thus, is subject to measurement errors, misreporting, and social desirability biases.^23^ Secondly, due to the lack of reliable data, this study did not explore the nature of symptoms that the respondents delayed or avoided seeking care. Thirdly, the study did not collect specifically any information on virtual consultations. Due to the risk involved with in-person visits during the pandemic, many respondents might have opted for teleconsultations. Despite these limitations, our study contributes to the limited evidence on the patterns of access to care during the pandemic. To the best of our knowledge, this is the first study that explores access to care, predictors and reasons during the COVID-19 pandemic in a nationally representative population of older adults.

## 5. Conclusion

Using the nationally representative HRS data, we found the older population in the USA to have reduced access to doctor’s visits, surgery, prescription filling, and dental care during the pandemic. The odds of reduced access were more among females, younger, educated, not receiving social security benefits, with limitations in daily activities, and those with three preexisting conditions. We suggest improving access to care for older adults by strengthening the availability of telehealth services and the involvement of medical volunteers and voluntary organizations. Awareness on the benefits of telehealth and the risks of delaying and neglecting care need to be reemphasized.

## Data Availability

The data used to support the findings of this study are available publicly through the Health and Retirement Study.

## Acknowledgements

We are grateful to the Health and Retirement Study for making this data publicly available.

